# Using Large Language Models for sentiment analysis of health-related social media data: empirical evaluation and practical tips

**DOI:** 10.1101/2024.03.19.24304544

**Authors:** Lu He, Samaneh Omranian, Susan McRoy, Kai Zheng

## Abstract

Health-related social media data generated by patients and the public provide valuable insights into patient experiences and opinions toward health issues such as vaccination and medical treatments. Using Natural Language Processing (NLP) methods to analyze such data, however, often requires high-quality annotations that are difficult to obtain. The recent emergence of Large Language Models (LLMs) such as the Generative Pre-trained Transformers (GPTs) has shown promising performance on a variety of NLP tasks in the health domain with little to no annotated data. However, their potential in analyzing health-related social media data remains underexplored. In this paper, we report empirical evaluations of LLMs (GPT-3.5-Turbo, FLAN-T5, and BERT-based models) on a common NLP task of health-related social media data: sentiment analysis for identifying opinions toward health issues. We explored how different prompting and fine-tuning strategies affect the performance of LLMs on social media datasets across diverse health topics, including Healthcare Reform, vaccination, mask wearing, and healthcare service quality. We found that LLMs outperformed VADER, a widely used off-the-shelf sentiment analysis tool, but are far from being able to produce accurate sentiment labels. However, their performance can be improved by data-specific prompts with information about the context, task, and targets. The highest performing LLMs are BERT-based models that were fine-tuned on aggregated data. We provided practical tips for researchers to use LLMs on health-related social media data for optimal outcomes. We also discuss future work needed to continue to improve the performance of LLMs for analyzing health-related social media data with minimal annotations.

## Introduction

Patients, caregivers, and the public produce rich content related to health on social media platforms. Extensive research has analyzed health-related social media data to gain insights into important health issues such as healthcare service quality, medical treatments and interventions, and healthcare policies^**1–3**^. Due to the large scale of health-related social media data, Natural Language Processing (NLP) techniques such as sentiment analysis and topic modeling prove to be valuable in this line of research. Sentiment analysis can be used in place of survey questionnaires to quickly obtain information about the uptake or rejection of important public health efforts (such as promotion of vaccines or masking), or how the public views particular policy initiatives such as Unites States health care reform. Related comments for all of these have been mentioned extensively in Tweets, and samples have been manually annotated for sentiment by experts, to create datasets (e.g. the Health Care Reform (HCR) dataset). Assessing public opinion in this way simplifies the process of repeated sampling during a rapidly evolving situation. However, accurate NLP techniques often require high-quality annotated data, which is particularly challenging to obtain in the health domain due to the variety of health topics discussed and the heterogeneity of these topics. Off-the-shelf tools, on the other hand, require no annotated data but suffer from low performances when applied in health contexts due to domain incompatibility. Our prior work found that most such studies have inconsistent and poor research design and reporting practices, which remains a significant concern around the validity and reliability of NLP-based analysis of health-related social media data^**4,5**^.

The recent emergence of Large Language Models (LLMs) brings excitement and new opportunities to NLP and the health domain. LLMs are pre-trained on gigantic corpora such as Wikipedia articles to learn and memorize semantic representations and relationships of natural language and are further tuned on various downstream NLP tasks, such as sentiment analysis or named entity recognition (NER) to enhance their capabilities^**6**^. Representative LLMs include the Bi-directional Encoder-Decoder Recurrent Transformer (BERT), Generative Pre-Trained LLM (GPT) by Open-AI, FLAN-T5 by Google, and LLaMA by Meta. These LLMs have shown superior performance and versatility in a variety of NLP tasks, such as question answering, NER, text summarization, and language translation. Researchers have set out to evaluate the capabilities of the LLMs on diverse datasets and tasks. For example, Ziems et al. evaluated LLMs from the GPT family and FLAN-T5 on multiple computational social science tasks such as toxicity detection and positive framing^**7**^. These studies can involve either so-called “zero-shot” evaluations, where only unlabeled test data is provided for analysis or “few-shot” tests or “fine-tuning”, where a sample of annotated data is provided for analysis prior to the presentation of test cases.

In the health domain, most recent evaluations of LLMs focus on clinical data such as clinical notes, Electronic Health Records (EHR), and medical images^**8**^. For example, Hu et al. developed multiple prompting strategies to evaluate the capabilities of zero-shot LLMs such as GPT-3.5 and GPT-4 on classic clinical NER tasks and found that, although they have lower performance compared to fine-tuning BioClinicalBERT on clinical data, they have demonstrated promising results with no annotated data required^**9**^. Goel et al. used LLMs as additional annotators in clinical NER tasks and found that they can lower clinical annotators’ burden while maintaining high performance of the NLP models^**10**^. Despite these promising results in evaluating LLMs on clinical NLP tasks, few assessments considered consumer health data. For example, Lossio-Ventura et al. evaluated zero-shot ChatGPT for analyzing the sentiments conveyed through COVID-19 free-text survey responses. With no annotated data provided, ChatGPT achieved an F-measure of 0.8668, outperforming many commonly used off-the-shelf tools including VADER and the Linguistic Inquiry and Word Count (LIWC)^**11**^. Fu et al., evaluated ChatGPT on patient-generated texts in Cantonese through online counseling sessions and found that the zero-shot LLM can identify sentiments with over 90% accuracy, outperforming multiple machine learning models developed with annotated datasets^**12**^. Deiner et al. evaluated GPTs’ capacity in rating the probabilities of tweets that indicate conjunctive outbreaks and found that GPT-4 had satisfying agreements with two human raters with Pearson correlation scores of 0.73 and 0.77^**13**^.

Despite these promising results suggesting that LLMs may change the landscape of NLP in the health domain, there is limited work that investigates their capacity to analyze the sentiments conveyed through health-related social media data. This type of data presents unique challenges such as the abundance of informal expressions created by patients and the public, and the constantly evolving languages that change in response to social media platform policies ^**14**^ or health crises^**2**^. Analyzing the sentiments and opinions from patient and public-generated health social media data remains a critical topic, as it provides valuable insights into patient and public concerns toward health issues and enables patient-centered care by incorporating their voices. Despite the critical need to analyze large-scale health social media data, research shows that existing computational tools were not compatible with social media data and health contexts. Our previous studies found that commonly used off-the-shelf tools performed extremely poorly when analyzing the sentiments of health-related social media data. Developing machine and deep learning models on study data also require high-quality annotated data which can be challenging to obtain.

To address the critical need of assessing the feasibility of utilizing LLMs in analyzing health-related social media data, we conducted a study to evaluate the performance of zero-shot and few-shot LLMs against LLMs fine-tuned on annotated data on various health-related social media datasets, covering vaccination for human papilloma virus (HPV), mask wearing during COVID-19, the US healthcare reform in the Obama era, and reviews of healthcare providers. A commonly used off-the-shelf tool was used as baseline for comparison. We varied prompting and fine-tuning strategies for the LLMs such as by incorporating aspect information in prompts and by assembling annotated data from multiple datasets for fine-tuning. We found that zero-shot and few-shot LLMs, though still far from satisfying, generally have better performance than the off-the-shelf tool. Incorporating data-specific information into the prompts improved model performance by a large margin on most of the datasets. BERT-based models that were fine-tuned on ensembled data showed the highest performance. Finally, we discuss the role of LLMs in analyzing patient and public-generated health social media data as well as practical tips for health informatics researchers to leverage them with little annotated data for better performance.

## Methods

### Evaluation datasets

We used datasets that were evaluated in our previous work^4^. The datasets were selected based on our systematic review and must include human coders’ annotation as ground truth labels.^5^ More detailed description of the datasets can be found in He et al.^4^ The evaluation datasets included four health-related social media datasets: Health Care Reform (“HCR”), Human Papillomavirus (“HPV”) Vaccine, COVID-19 Masking (“Mask”), and Vitals.com Physician Reviews (“Vitals”). A non-health dataset, the IMDB Dataset (“IMDB”) created from movie reviews, was used for baseline comparison. To reduce the cost of OpenAI API usage, we randomly sampled 1,000 posts from the Vitals dataset and the IMDB dataset, respectively.

The HCR dataset was originally created by Speriosu et al. to study the public’s reactions to the Health Care Reform in the Obama era^15^. The dataset includes 2,315 tweets, with 701 labeled as positive, 1,034 as negative, and 580 neutral. The HPV dataset was released by Du et al. for identifying public’s opinions toward the HPV vaccination^16^. The data consists of 964 positive, 1,044 negative, and 1,085 neutral tweets. The Mask dataset was collected, labeled, and maintained by our research team to study public attitudes and concerns toward mask wearing during the COVID-19 pandemic.^2^ The dataset was collected from Twitter between January 1 and November 1, 2020. A random sample of 609 tweets were annotated; 530 tweets supported mask wearing and 79 opposed mask wearing. The Vitals data was collected from one of the largest online physician review websites, Vitals.com, where patients and caregivers wrote reviews to rate the quality of healthcare services they received^3^. The sentiment labels were determined based on the numeric rating provided by the users (e.g., reviews with 5-star were labeled as positive and reviews with 1-star were negative). In the 1,000 random samples used in this study, 744 are positive reviews and 256 are negative reviews. The IMDB dataset was released by Maas et al., which included movie reviews collected from one of the largest movie review websites, IMDB.com, is a commonly used benchmark dataset for sentiment analysis and is used as a comparison dataset in this study^17^. In the 1,000 random samples included in this study, 623 are negative movie reviews and 377 are positive.

### Models and tools selected for evaluation

#### Zero-shot LLMs

We selected widely used LLMs that can generate textual responses to zero-shot NLP tasks with natural language prompts. Based on recent studies that evaluated LLMs in the general domain and on clinical notes ^7,9,18,19^, we selected FLAN-T5 by Google ^20^ and GPT-3.5-Turbo^21^ by OpenAI. We initially also experimented with LLaMA2 provided by Meta, but it failed to produce answers in consistent formats and sometimes merely repeated the question prompts. Therefore, we did not include LLaMA2 in this evaluation study. We accessed GPT-3.5-Turbo through OpenAI’s Chat Completion API^22^. FLAN-T5 was accessed via Huggingface Transformer API^23^. For all experiments involving GPT, the temperature was set to 0 to reduce randomness and ensure the consistency of outputs from the models.

Extensively experimenting with all checkpoints of the LLMs is beyond the scope and capacity of this study. We instead focused on the commonly used versions of the LLMs. The GPT-3.5-Turbo-1106 checkpoint and FLAN-T5-Base were used.^20^ The FLAN-T5-Base checkpoint is a fine-tuned T5 model optimized for more than 1000 downstream tasks and contains more than 250 million parameters.

#### Fine-tuned LLMs

We also selected commonly used pre-trained LLMs and fine-tuned them on a held-out set of our evaluation datasets. We used a general LLM (BERT_base_uncased) and domain and task-specific LLMs (BioClinicalBERT^25^ and SiEBERT^26^). BERT-based models have been widely used in downstream NLP tasks such as text classification and named entity extraction^27^. We accessed the pre-trained models through the HuggingFace model repository and fine-tuned the models using the Trainer API. It is beyond the scope of this study to comprehensively evaluate all available pre-trained LLMs. We only selected commonly used models that are relevant to the task of sentiment analysis and the health domain.

#### Baseline off-the-shelf tools

We used Valence Aware Dictionary and sEntiment Reasoner (VADER) as the baseline off-the-shelf tool for comparison, as it produced the highest performance in an evaluation study on the same datasets^4^. VADER is an open-source Python package using lexicons and manually curated rules to classify sentiments of social media data^28^. It computes a sentiment score on a piece of input text. Based on a threshold provided in the package, a sentiment label will be produced. More information about the tool and its performance is documented in He et al^4^.

### Experiments and setups

#### Experiment 1: comparing zero-shot LLMs with fine-tuned LLMs and the baseline off-the-shelf tool

The goal of Experiment 1 was to evaluate zero-shot LLMs against fine-tuned LLMs on the evaluation datasets with VADER being the baseline. In this experiment, zero-shot LLMs received no annotation data. Natural language prompts were curated moderately so that the LLMs can produce usable and comprehensible results, without any specific references to the data or the context of the data. The zero-shot LLMs were evaluated on full datasets because no training or validation data were needed. The finalized prompt used for all zero-shot LLMs across all datasets is: “*What is the sentiment of the text? Only return the letter representing the answer. A. positive B. negative C. neutral D. not sure Text: [REPLACE WITH DATA]*.”

Pre-trained BERT-based LLMs (BERT_base_uncased and BioClinicalBERT) were fine-tuned on a random sample of the evaluation datasets. Each evaluation dataset was split into a training, validation, and test set, following a 0.8-0.1-0.1 ratio. SiEBERT (Sentiment in English) was already fine-tuned on various texts with sentiment labels, including reviews and tweets^26^. Therefore, we directly adopted the fine-tuned check point of the model, without further tuning using our study data. Since the maximum token size allowed for BERT-based models is 512, we truncated the input text to include only 512 tokens; texts containing less than 512 tokens were padded with extra tokens so that input texts are of the same length.

#### Experiment 2: comparing zero-shot LLMs using generic versus data-specific prompts

The goal of Experiment 2 was to investigate whether and how different prompting strategies affect the performance of the zero-shot LLMs. Subsequently, we tested basic prompts against data-specific prompts. We also tested chain-of-thought prompts which required LLMs to reason and have shown promising results in prior studies^29^. However, the performance from our pilot study was extremely poor when using chain-of-thought prompts and even worse than generic prompts. This is probably because chain-of-thought prompts are useful for simple reasoning tasks such as solving mathematical problems or answering questions. Therefore, we decided to exclude chain-of-thought prompts in our evaluation.

While there are many viable prompts, to make a fair baseline for the basic and generic prompts, we only conducted moderate prompt engineering, following instructions provided in previous studies^7^. Because sentiment analysis is essentially a text classification task with pre-defined sentiment labels, we formatted the prompts as multiple-choice problems, where each option is listed after a letter, followed by a new line character. We also instructed the LLMs to only produce the letter, both to reduce the number of tokens generated and to make the evaluation easier. After rounds of testing, we finalized the generic prompt and used it across all LLMs and on all evaluation datasets. To ensure that LLMs do not produce hallucinations on data that they are not confident in, we added another choice “Unsure”.

For designing data-specific prompts, we injected data-specific information into the generic prompt by providing the task information (classification of attitude as supporting or opposing, or classification of overall sentiment as positive and negative), target (mask, HPV, HCR, movie, provider), data (tweet, movie review, online physician review), and context if applicable (e.g., during the pandemic, Healthcare Reform by Obama). Each evaluation dataset has its own data-specific prompt. For example, the data-specific prompt for the Mask dataset is: “Does the tweet support mask wearing during the pandemic?” The data-specific prompts used for the datasets are shown in Figure 1 below.

**Figure 1.**
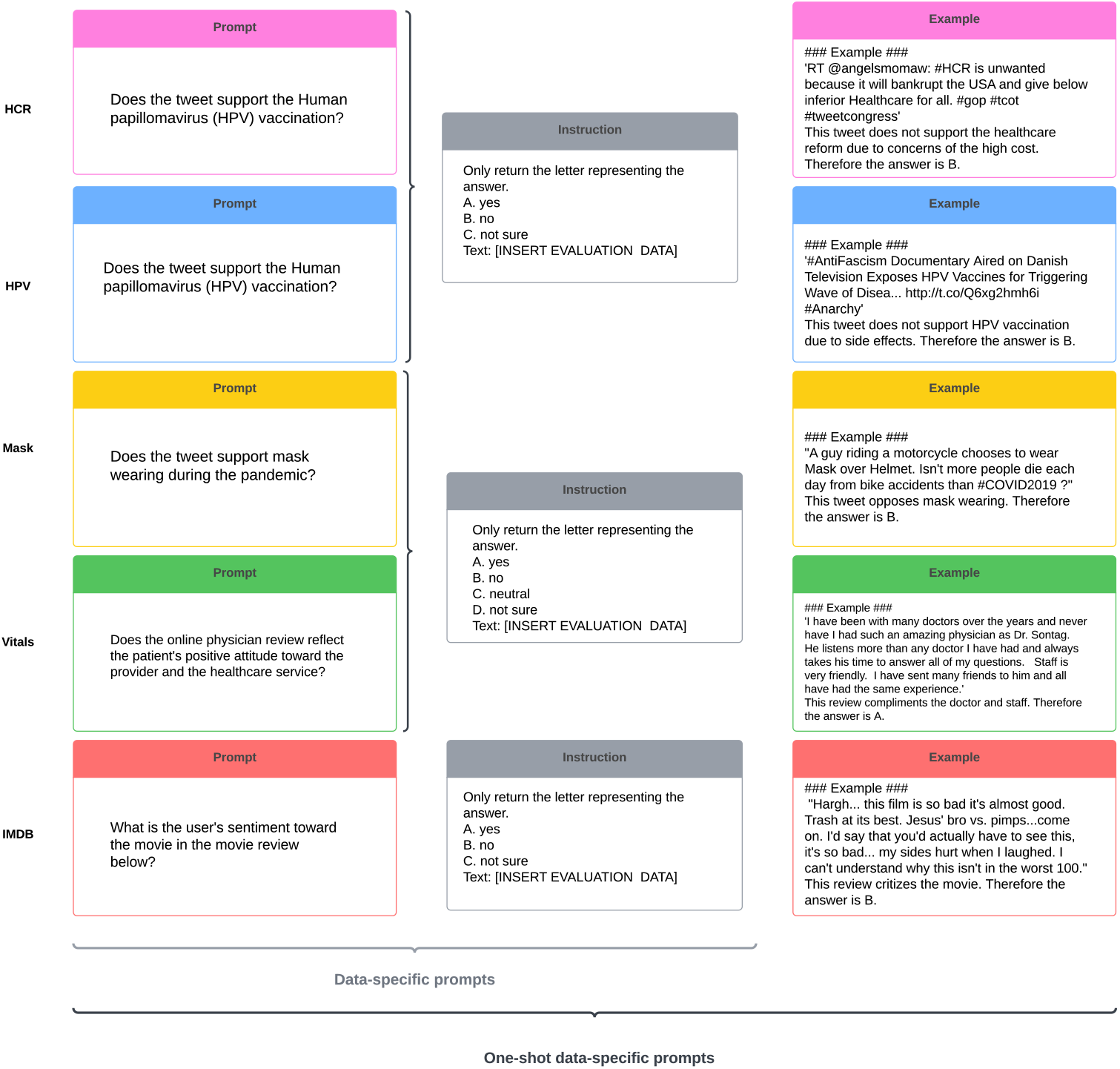
Data-specific prompts in zero and one-shot experiments.

#### Experiment 3: comparing zero-shot and one-shot LLMs

The goal of Experiment 3 was to investigate whether providing LLMs with annotated examples can outperform zero-shot LLMs, where no annotated example was provided. As it is not our focus to comprehensively assess the differences between the number of examples, we evaluated one-shot LLMs to control the cost of using OpenAI API. We extended each data-specific prompt by adding an example post from the data, along with a short explanation and the answer. The prompts used for one-shot LLMs are shown in Figure 1.

#### Experiment 4: comparing LLMs fine-tuned on single datasets versus models fine-tuned on an ensembled dataset

The goal of Experiment 4 was to investigate whether fine-tuning LLMs on ensembled datasets from multiple sources outperform LLMs fine-tuned on single-source datasets. To create the ensembled dataset, we aggregated all data from the five evaluation datasets and then randomly partitioned the ensembled dataset into training, validation, and test sets following the 0.8-0.1-0.1 ratio. The BERT-based models were fine-tuned on the training and validation sets and evaluated on individual test sets from each evaluation dataset so that the performance is a fair comparison with results from the BERT-based models fine-tuned on individual datasets. For example, BERT_base_ensemble was fine-tuned on a random set of 80% of all data aggregated from the five evaluation datasets. It was then evaluated individually on random sets of 10% of each of the five datasets.

## Results

### Experiment 1: Performance comparison between LLMs and off-the-shelf tools

Table 1 shows the weighted F1-scores of the zero-shot LLMs with generic prompts on all datasets, fine-tuned LLMs evaluated on sampled test sets of the data, and VADER on all data. Even without substantial prompt engineering or annotated examples to the zero-shot LLMs, GPT3.5-Turbo outperformed VADER on four out of five datasets and FLAN-T5 outperformed VADER on three out of five datasets.

**Table 1.** Weighted F1-scores of the models on the evaluation datasets. GPT-3.5-Turbo and FLAN-T5 are all zero-shot. BERT-based models are evaluated on held-out test sets. Highest weighted F1-scores are shown **bolded**.

|  | Zero-shot LLMs |  | Fine-tuned LLMs |  |  | Baseline tool |
| --- | --- | --- | --- | --- | --- | --- |
|  | GPT3.5-Turbo | FLAN-T5 | BERT_base | SiEBERT | BioClinicalBERT | VADER |
| HCR | 0.57 | 0.40 | 0.52 | **0.60** | 0.57 | 0.41 |
| HPV | 0.55 | 0.34 | **0.59** | 0.39 | 0.51 | 0.49 |
| Mask | 0.22 | 0.64 | 0.48 | 0.58 | **0.73** | 0.59 |
| Vitals | 0.95 | **0.97** | 0.64 | **0.97** | 0.68 | 0.89 |
| IMDB | 0.75 | 0.94 | 0.21 | **0.95** | 0.45 | 0.65 |

Though zero-shot LLMs have demonstrated higher performance than VADER, their results are still far from usable for research on most of the datasets, with weighted F1-scores lower than 0.6. The LLMs achieved the highest performance on the Vitals dataset, with a weighted F1-score of 0.97. Notably, both GPT3.5-Turbo and FLAN-T5 have extremely poor performance on the HPV and HCR data. The three twitter datasets (HCR, HPV, and Mask) are still challenging for the LLMs models, despite the improved performance over the baseline, VADER.

Out of the six tools/LLMs evaluated, SiEBERT performed the best on three out of five datasets, suggesting that LLMs fine-tuned on large scale datasets can indeed generalize to new datasets, though their applicability to health-related social media data such as HPV and Mask is still limited.

### Experiment 2 & 3: Performance comparison between LLMs using basic and data-specific prompts in zero and one-shot settings

Table 2 shows weighted F1-scores based on results from zero and one-shot LLMs using generic prompts and data-specific prompts. For zero-shot GPT-3.5-Turbo, the performance increased by large margins when using data-specific prompts on three out of the five datasets. However, it had a lower performance with data-specific prompts on the HPV dataset. The Mask and IMDB datasets benefited the most from using data-specific prompts. While GPT-3.5-Turbo generally benefited from data-specific prompts, FLAN-T5 produced much poorer results when data-specific prompts were used, except for on the Mask dataset. For both the HCR and HPV datasets, none of the models performed better with data-specific prompts, although GPT-3.5-Turbo saw slight improvement on the HCR data in the data-specific, one-shot condition.

**Table 2.**
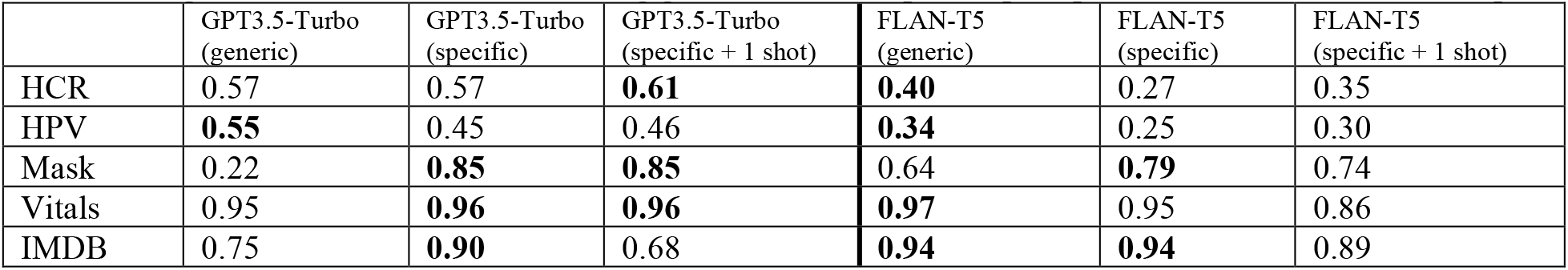
Weighted F1-scores of the LLMs using generic and data-specific prompts and with an annotated example.

|  | GPT3.5-Turbo (generic) | GPT3.5-Turbo (specific) | GPT3.5-Turbo (specific + 1 shot) | FLAN-T5 (generic) | FLAN-T5 (specific) | FLAN-T5 (specific + 1 shot) |
| --- | --- | --- | --- | --- | --- | --- |
| HCR | 0.57 | 0.57 | **0.61** | **0.40** | 0.27 | 0.35 |
| HPV | **0.55** | 0.45 | 0.46 | **0.34** | 0.25 | 0.30 |
| Mask | 0.22 | **0.85** | **0.85** | 0.64 | **0.79** | 0.74 |
| Vitals | 0.95 | **0.96** | **0.96** | **0.97** | 0.95 | 0.86 |
| IMDB | 0.75 | **0.90** | 0.68 | **0.94** | **0.94** | 0.89 |

Across the datasets, the Mask dataset gained the most from data-specific prompts. This is in part attributable to the importance of the context in which the Mask dataset was created. Without the contexts of the aspect (mask wearing) and during the pandemic, it is even challenging for human annotators to label the attitudes.

Comparing the LLMs in zero and one-shot settings, we did not observe benefits from the addition of an annotated example for GPT-3.5-Turbo and FLAN-T5. For FLAN-T5, the performance even decreased on three datasets (Mask, Vitals, IMDB). For GPT3.5-Turbo, adding an annotated example marginally improved the performance on the HCR and HPV dataset, Overall, data-specific prompts improved performance for GPT3.5-Turbo but not for FLAN-T5. The benefits from adding an annotated example were not observed for both models.

### Experiment 4: Performance comparison between fine-tuning LLMs on individual datasets versus an ensembled dataset

**Table 3** below compares the weighted F1-scores across the BERT-based models that were fine-tuned on individual test sets with those fine-tuned on an ensembled test set. In general, those tuned on the ensembled set outperformed by a large margin. Even the most basic BERT model fine-tuned on ensembled data achieved the highest weighted F1-scores. Notably, the BERT_base_ensemble also achieved the highest performance on the HPV datasets among all models, with a weighted F1-score of 0.94, whereas the performance of other models was less than 0.6. This finding is not surprising, given that more and diverse annotated data for fine-tuning can usually improve model performance. The large margin of improvement, however, suggests that annotated data from other contexts are still useful for downstream tasks in different contexts. Notably, the performance achieved by the BERT-based models fine-tuned on the ensembled data is also the highest among all tools and models. For the HCR, HPV, and Mask data, the fine-tuned BERT-based models even achieved weighted F1-scores higher than 0.85. BERT_base_ensemble consistently outperformed BioClinicalBERT_ensemble, possibly because BERT_base was pre-trained on general English corpora that aligns better with social media data, while BioClinicalBERT was pre-trained on clinical notes and the underlying semantic representations are quite different.

**Table 3.**
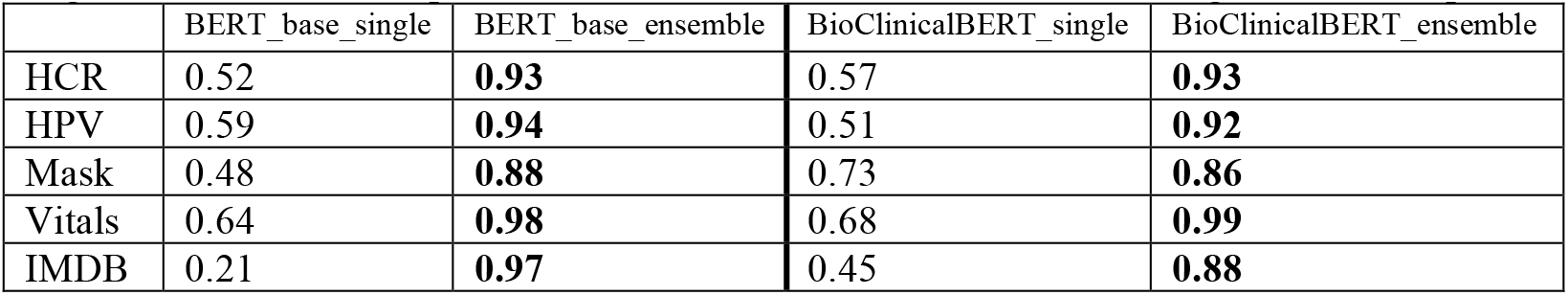
Weighted F1-scores of the pre-trained, BERT-based models fine-tuned using different samples.

|  | BERT_base_single | BERT_base_ensemble | BioClinicalBERT_single | BioClinicalBERT_ensemble |
| --- | --- | --- | --- | --- |
| HCR | 0.52 | **0.93** | 0.57 | **0.93** |
| HPV | 0.59 | **0.94** | 0.51 | **0.92** |
| Mask | 0.48 | **0.88** | 0.73 | **0.86** |
| Vitals | 0.64 | **0.98** | 0.68 | **0.99** |
| IMDB | 0.21 | **0.97** | 0.45 | **0.88** |

## Discussion

### Overall performance and observations from the experiments

Our evaluation results show that while LLMs demonstrated promising performance on a variety of NLP tasks in other domains and on clinical data, they are still far from reliable when applied on health-related social media data, especially Twitter data that are known to be challenging for NLP. The performance of LLMs highly depends on the nature of the health-related social media data and how researchers formulate the tasks and design the prompts. Online physician reviews are less challenging for LLMs, as they most directly express sentiments and opinions. The performance of zero-shot LLMs on the Vitals dataset is above 0.95 for GPT-3.5-Turbo and FLAN-T5, with no annotated data provided or prompting strategies used. However, on the more challenging datasets such as tweets related to HCR, HPV, and mask wearing, the highest weighted F1-score is only slightly above 0.6, achieved by FLAN-T5-base on the Mask dataset.

The datasets and models also responded differently to further prompt tuning. Embedding data-specific information into prompts may enhance the performance of zero-shot LLMs, but the benefits depend on the data and contexts. For example, the Mask dataset benefited the most from data-specific prompts, but the HCR and HPV datasets did not. FLAN-T5 saw reduced performance when data-specific prompts were used. Adding an annotated example also did not improve the performance of GPT-3.5-Turbo and FLAN-T5.

The highest performing models from our evaluation study are BERT-based models fine-tuned on an ensembled dataset. The fine-tuned models achieved weighted F1-scores higher than 0.85, even on the more challenging datasets such as the HCR, HPV, and Mask dataset. The basic BERT model consistently outperformed BioClinicalBERT, possibly due to the alignment between health social media data and the generic English corpora that BERT was pre-trained on. Surprisingly, BioClinicalBERT that was pre-trained on clinical notes from Intensive Care Unit settings still showed good performance, despite the development domain highly differing from the social media contexts. This may suggest that having a diverse ensembled data for further fine-tuning can still improve model performance and efficiently transfer the previously learned semantic representations to a new domain.

Overall, our evaluation results suggest that GPT and FLAN-T5 are still not yet directly usable for analyzing health-related social media data, especially data related to healthcare policy and vaccination. Using data-specific prompts can improve the performance of the models on some datasets but not consistently. For more reliable results, researchers should still leverage annotated data to fine-tune pre-trained models such as BERT and BioClinicalBERT. Assembling diverse data for fine-tuning can greatly improve the performance of these models.

### Practical tips for using LLMs on health-related social media data

While the model behaviors and performances are difficult to analyze, we provide a set of practical tips derived from the series of experiments we conducted. However, it is worth noting that the tips may only be applicable to certain social media and health contexts. Researchers should explore their study data and investigate what works the best for their specific study contexts.

First, correctly formulating the sentiment analysis task and providing this information to LLMs is essential for better performance. While sentiment analysis generally refers to the task of identifying sentiments, opinions, and attitudes from texts, it can be manifested very differently in different study contexts. For example, on the Mask dataset, the task is formulated as “identifying the attitudes as supporting or opposing” (a form of argumentation), while on the Vitals dataset, the task is “identifying the emotion toward the provider as positive or negative”.

Second, health-related social media data often contain multiple aspects at the same time, therefore, specifying the intended focus where sentiment is wanted is important for the LLMs. For example, in the Mask data, users commonly used negative words to express their supporting attitudes toward mask wearing. This, however, often requires researchers to first conduct exploratory qualitative review of their data to identify aspects of interest to correctly instruct the LLMs.

Third, while LLMs are all pre-trained on gigantic corpora and have a huge parameter space, their underlying architecture and therefore their specialties still differ. For example, according to Ziems et al., FLAN-T5 performed better than GPT models on sentiment classification on most of the datasets^7^. Our evaluation results also found that the models responded differently to data-specific prompts and the additional annotated examples. This suggests that researchers should familiarize themselves with the unique features of the LLMs to find the model that best fits their study data and contexts. In general, we observed that FLAN-T5 works best with simple and straightforward prompts, while GPT models benefit from detailed natural language prompts with information about study data and contexts.

### The role of zero and few-shot learning LLMs in analyzing health-related social media text

While our evaluation results show that zero-shot LLMs are still far from capable of producing reliable sentiment labels on health-related social media data, they have several strengths that make them good candidates for being incorporated into the analysis workflow. First, researchers can interact with zero or few-shot LLMs such as GPT and FLAN-T5 through natural language-based prompts, lowering the technical requirements of training, developing, and fine-tuning their own deep learning models. In addition, researchers can easily extend the analysis to more granular levels, such as aspect-based sentiment analysis, with zero-shot LLMs. Analyzing patient and public-generated social media data may particularly benefit from the power and promises of zero and few-shot LLMs. Unlike clinical NLP tasks where the clinical information and entities of interest are relatively fixed, health-related social media data are often heterogeneous and evolving. The nature of health-related social media data often requires researchers to conduct manual review at the beginning to identify aspects of interest. Zero and few-shot LLMs and their versatility and flexibility can potentially help researchers to conduct exploratory analysis of health-related social media data, especially when the content of the data is not clear. In the process, researchers can also improve or tune the model using natural language input, leading to efficient synergies between LLMs and researchers.

While researchers can be freed from tedious data annotation, feature engineering, model development and validation, their roles may be as critical in the LLM era. For example, it is important for the LLMs to have accurate and relevant prompts for the tasks, especially the context, sentiment analysis target, and data. This requires researchers to have a good understanding of their study data and invest time to conduct exploratory analysis of the data.

Our results from Experiment 4 also shows that BERT-based LLMs performed the best when fine-tuned on ensembled data from all evaluation datasets. This, however, requires a larger amount of annotated data for fine-tuning. A possible workaround is to use the zero-shot LLMs to generate synthetic data with labels and use the generated data to augment the fine-tuning dataset with BERT-based models. Though zero or few-shot LLMs themselves may not be able to produce accurate sentiment labels, their superior ability to generate human-like texts can be leveraged in augmenting annotated data to improve the performance on downstream tasks.

### Challenges and issues with using LLMs in analyzing patient and public-generated social media data

Despite the promising performance and the ease of use of the zero-shot LLMs on health-related social media data, we also note challenges and issues that researchers should be aware of when using these LLMs. First, most of the LLMs are proprietary products made available through commercial companies such as OpenAI, Google, and Meta. While FLAN-T5 by Google and Llama2 by Meta are currently freely available or upon request, it is hard to predict whether and at what price they will charge for using their models. The rising costs of close-sourced LLMs such as GPTs also raise concerns about the feasibility of large-scale analysis of social media data. Currently, it costs $10 for 1 million tokens with GPT-4-Turbo, which is 200 times the price for GPT-3-Turbo. It is unknown whether they will continue to increase and what consequences this may have on researchers with limited financial resources.

Our results also show that there are no consistent patterns of which LLM and what prompts work the best for what types of data. OpenAI’s models are not open-sourced, making the use and explanation of the model output opaque for researchers in the health domain. Second, recent research has revealed the risk of LLMs in leaking training data, which could be particularly concerning due to the sensitive nature of health-related data.^30^ While OpenAI stated that user input will only be stored on their servers and not be used for training their models, patients and the public who produced the health-related social media data may still perceive this as risky. The recent deal between Reddit, one of the largest online communities, and an unknown company to access all public Reddit data for developing LLMs also raised concerns of how public social media data will be used in the LLM era.^31^ This could be particularly concerning for patients who produce health-related content on social media platforms, as their posts may contain personal and sensitive information. This warrants future studies to investigate how to best protect patient and public-generated social media data when LLMs are trained on and used for analyzing them.

### Limitations

Our study is limited to the selection of health-related social media data that are publicly accessible and have been annotated by experts. While we tried to diversify the health topics covered and social media platforms studied, three out of the five datasets we evaluated were from Twitter data. In addition, due to the cost of using the OpenAI API, we were not able to comprehensively experiment with an exhaustive set of prompting strategies or model checkpoints. The number of available LLMs is also increasing rapidly; it is beyond the scope and capacity of this work to compare all available LLMs on the study data.

### Future directions

This evaluation study sets the stage for exciting future work. First, we will further explore how LLM sizes affect their performance, ie, whether larger LLMs always have better performance or whether more easily distributed smaller models would suffice. Second, we will leverage the retrieval-augmented generation (RAG) framework with a small set of annotated health-related social media data to improve the LLMs and further reduce the risk of hallucination. We will also explore innovative use of LLMs to improve the performance, such as by using LLMs to generate synthetic data to augment the size of training data and improve downstream tasks. Third, we will further evaluate the capabilities of zero-shot LLMs on more NLP tasks such as topic modeling and misinformation detection using health-related social media data. Last, we will evaluate the capabilities of LLMs on multimodal health-related social media data, combining texts, images, audios, and videos for a more comprehensive analysis of patient and public-generated data.

## Conclusions

LLMs such as GPT-3.5-Turbo and FLAN-T5 consistently perform better than a highly regarded, rule-based off-the-shelf tool when analyzing the sentiments and opinions of health-related social media data, even when no annotated data is provided. Further, enriching prompts with data-specific details about targets of the opinions and social media platforms improves the performance of current LLMs on most of the evaluation datasets. This evaluation study shows the potential of using LLMs in computational analysis of health-related social media data with little to no annotated data. However, researchers will still need to have a good understanding of the study data they are working with, to correctly instruct LLMs to produce accurate results. Future work should further investigate how to continuously tune LLMs with domain data for better performance.

## Data Availability

Data will be shared upon reasonable request.

## Notes

### Competing Interest Statement

The authors have declared no competing interest.

### Funding Statement

No funding.

## References

1. Davis MA, Zheng K, Liu Y, Levy H. Public Response to Obamacare on Twitter. J Med Internet Res. 2017;19(5). doi:10.2196/jmir.6946

2. He L, He C, Reynolds TL, et al. Why do people oppose mask wearing? A comprehensive analysis of U.S. tweets during the COVID-19 pandemic. J Am Med Inform Assoc. 2021;28(7):1564–1573. doi:10.1093/jamia/ocab047

3. He L, He C, Wang Y, Hu Z, Zheng K, Chen Y. What Do Patients Care About? Mining Fine-grained Patient Concerns from Online Physician Reviews Through Computer-Assisted Multi-level Qualitative Analysis. AMIA Annu Symp Proc. 2021;2020:544–553.

4. He L, Yin T, Zheng K. They May Not Work! An evaluation of eleven sentiment analysis tools on seven social media datasets. J Biomed Inform. 2022;132:104142. doi:10.1016/j.jbi.2022.104142

5. He L, Yin T, Hu Z, Chen Y, Hanauer DA, Zheng K. Developing a standardized protocol for computational sentiment analysis research using health-related social media data. J Am Med Inform Assoc JAMIA. 2020;28(6):1125–1134. doi:10.1093/jamia/ocaa298

6. Brown TB, Mann B, Ryder N, et al. Language Models are Few-Shot Learners. Published online July 22, 2020. doi:10.48550/arXiv.2005.14165

7. Ziems C, Held W, Shaikh O, Chen J, Zhang Z, Yang D. Can Large Language Models Transform Computational Social Science? Published online December 7, 2023. doi:10.48550/arXiv.2305.03514

8. Tian S, Jin Q, Yeganova L, et al. Opportunities and challenges for ChatGPT and large language models in biomedicine and health. Brief Bioinform. 2024;25(1):bbad493. doi:10.1093/bib/bbad493

9. Hu Y, Chen Q, Du J, et al. Improving large language models for clinical named entity recognition via prompt engineering. J Am Med Inform Assoc. Published online January 27, 2024:ocad259. doi:10.1093/jamia/ocad259

10. Goel A, Gueta A, Gilon O, et al. LLMs Accelerate Annotation for Medical Information Extraction. In: Proceedings of the 3rd Machine Learning for Health Symposium. PMLR; 2023:82–100. Accessed December 15, 2023. https://proceedings.mlr.press/v225/goel23a.html

11. Lossio-Ventura JA, Weger R, Lee AY, et al. A Comparison of ChatGPT and Fine-Tuned Open Pre-Trained Transformers (OPT) Against Widely Used Sentiment Analysis Tools: Sentiment Analysis of COVID-19 Survey Data. JMIR Ment Health. 2024;11(1):e50150. doi:10.2196/50150

12. Fu Z, Hsu YC, Chan CS, Lau CM, Liu J, Yip PSF. Efficacy of ChatGPT in Cantonese Sentiment Analysis: Comparative Study. J Med Internet Res. 2024;26(1):e51069. doi:10.2196/51069

13. Deiner MS, Deiner NA, Hristidis V, et al. Use of Large Language Models to Assess the Likelihood of Epidemics From the Content of Tweets: Infodemiology Study. J Med Internet Res. 2024;26(1):e49139. doi:10.2196/49139

14. Chancellor S, Pater JA, Clear TA, Gilbert E, De Choudhury M. #thyghgapp: Instagram Content Moderation and Lexical Variation in Pro-Eating Disorder Communities. In: Proceedings of the 19th ACM Conference on Computer-Supported Cooperative Work & Social Computing - CSCW ‘16. ACM Press; 2016:1199–1211. doi:10.1145/2818048.2819963

15. Speriosu M, Sudan N, Upadhyay S, Baldridge J. Twitter polarity classification with label propagation over lexical links and the follower graph. In: Proceedings of the First Workshop on Unsupervised Learning in NLP. EMNLP ‘11. Association for Computational Linguistics; 2011:53–63.

16. Du J, Xu J, Song H, Liu X, Tao C. Optimization on machine learning based approaches for sentiment analysis on HPV vaccines related tweets. J Biomed Semant. 2017;8. doi:10.1186/s13326-017-0120-6

17. Maas AL, Daly RE, Pham PT, Huang D, Ng AY, Potts C. Learning word vectors for sentiment analysis. In: Proceedings of the 49th Annual Meeting of the Association for Computational Linguistics: Human Language Technologies - Volume 1. HLT ‘11. Association for Computational Linguistics; 2011:142–150.

18. Ramachandran GK, Fu Y, Han B, et al. Prompt-based Extraction of Social Determinants of Health Using Few-shot Learning. In: Proceedings of the 5th Clinical Natural Language Processing Workshop. Association for Computational Linguistics; 2023:385–393. doi:10.18653/v1/2023.clinicalnlp-1.41

19. Guevara M, Chen S, Thomas S, et al. Large language models to identify social determinants of health in electronic health records. Npj Digit Med. 2024;7(1):1–14. doi:10.1038/s41746-023-00970-0

20. Chung HW, Hou L, Longpre S, et al. Scaling Instruction-Finetuned Language Models. Published online December 6, 2022. doi:10.48550/arXiv.2210.11416

21. Qin C, Zhang A, Zhang Z, Chen J, Yasunaga M, Yang D. Is ChatGPT a General-Purpose Natural Language Processing Task Solver? In: Bouamor H, Pino J, Bali K, eds. Proceedings of the 2023 Conference on Empirical Methods in Natural Language Processing. Association for Computational Linguistics; 2023:1339–1384. doi:10.18653/v1/2023.emnlp-main.85

22. OpenAI Platform. Accessed March 13, 2024. https://platform.openai.com

23. google/flan-t5-base · Hugging Face. Published January 4, 2024. Accessed March 13, 2024. https://huggingface.co/google/flan-t5-base

24. OpenAI, Achiam J, Adler S, et al. GPT-4 Technical Report. Published online March 4, 2024. doi:10.48550/arXiv.2303.08774

25. Alsentzer E, Murphy J, Boag W, et al. Publicly Available Clinical BERT Embeddings. In: Rumshisky A, Roberts K, Bethard S, Naumann T, eds. Proceedings of the 2nd Clinical Natural Language Processing Workshop. Association for Computational Linguistics; 2019:72–78. doi:10.18653/v1/W19-1909

26. Hartmann J, Heitmann M, Siebert C, Schamp C. More than a Feeling: Accuracy and Application of Sentiment Analysis. Int J Res Mark. 2023;40(1):75–87. doi:10.1016/j.ijresmar.2022.05.005

27. Si Y, Wang J, Xu H, Roberts K. Enhancing clinical concept extraction with contextual embeddings. J Am Med Inform Assoc. 2019;26(11):1297–1304. doi:10.1093/jamia/ocz096

28. Hutto CJ, Gilbert E. VADER: A Parsimonious Rule-Based Model for Sentiment Analysis of Social Media Text. In: Eighth International AAAI Conference on Weblogs and Social Media. ; 2014. Accessed July 15, 2019. https://www.aaai.org/ocs/index.php/ICWSM/ICWSM14/paper/view/8109

29. Wei J, Wang X, Schuurmans D, et al. Chain-of-Thought Prompting Elicits Reasoning in Large Language Models. Adv Neural Inf Process Syst. 2022;35:24824–24837.

30. Liu Y, Yan C, Yin Z, et al. Biomedical Research Cohort Membership Disclosure on Social Media. AMIA Annu Symp Proc. 2020;2019:607–616.

31. Reddit has a new AI training deal to sell user content - The Verge. Accessed March 15, 2024. https://www.theverge.com/2024/2/17/24075670/reddit-ai-training-license-deal-user-content

